# Differential effects of frailty on biventricular function and prognosis analysis in senile patients

**DOI:** 10.1101/2023.02.27.23286544

**Authors:** Jiang Jing, Yang Baojun, Zhiying Zhao, Xie Dili, Zhang Yunhe, Yu Rong, Jin Jing

## Abstract

**Objective:** To investigate the differential effects of frailty on biventricular function in senile patients and analyse the prognosis of different combinations of clinical status.

**Methods and Results:** Patients aged ≥80 years with at least one basic disease causing heart failure were included and divided into three groups according to frailty score. Basic data, ultrasound data, and follow-up data were collected and analyses of differences between groups and survival were performed. The proportion of patients with right heart failure in the frailty group was significantly higher than that in the others. A total of 33 (15.1%) patients died within a year, 162 (74%) were readmitted within 1 year, and 84 (38.4%) were admitted for heart failure within 1 year. The frailty group with right heart failure had the highest rate of all cause and heart failure-related readmission. Frailty significantly increased the risk of 1-year all-cause mortality, all-cause readmission, and heart failure-related readmission. Right heart failure significantly increased the 1-year all-cause readmission and heart failure-related readmission rates. After adjusting for the interaction of factors, only frailty had a significant effect on the three prognostic events.

**Conclusions:** Right heart failure is more likely to be associated with frailty in senile patients. One-year all-cause mortality, all-cause readmission, and heart failure-related readmission rates were significantly increased in frail patients with right heart failure.

Frailty was a significant predictor of all-cause death, all-cause readmission, and heart failure-related readmission.

## Introductions

Heart failure is the terminal form of various cardiovascular diseases. Owing to the progress of medical technology, mortality related to heart failure has decreased, while the morbidity has increased, which has brought enormous economic pressure on the healthcare sector worldwide.[1] Therefore, there is an urgent need to improve treatment and management and reduce the hospitalization rate of patients with heart failure.

As the most common geriatric syndrome, frailty has been garnering increasing attention in present geriatric society. Frailty has gradually been explored in the field of heart failure. Studies showed that frailty was six times more common in patients with heart failure [4] and was significantly associated with poor prognosis; admission and all-cause mortality rates for heart failure increased by 40% and 48%, respectively, when associated with frailty.[2]

Therefore, the importance of identifying frailty was self-evident. Back in 2016 the European Heart Association recommended frailty screening for patients with heart failure.[5] However, the manifestations of frailty were not specific. Although the reliability of multiple scales has been hotly discussed, there is still no accepted protocol for research or clinical application.[3]

However, what aspect of frailty affects heart failure or heart function? These two syndromes may share common pathophysiological pathways, such as cellular senescence, oxidative stress, abnormal autophagy, and deoxyribonucleic acid damage[7]. It has been reported that heart failure with preserved ejection fraction has a higher incidence in patients with frailty than heart failure with reduced ejection fraction.[6] In terms of whether frailty predominantly affects left or right heart function, there remain no available studies.

Although the current research on the treatment of heart failure is mature, most studies focus on left ventricular function, especially left ventricular systolic function.[8] For right ventricular dysfunction, both clinical manifestations and treatment plan differ from those of left ventricular dysfunction. Therefore, our study focused on the different effects of frailty on biventricular function and prognosis to provide a basis for future treatment of frailty in patients with heart failure.

## Methods

### Study Population

From January 2020 to December 2020, 657 patients aged ≥80 years old were admitted to the Geriatric Department of Sichuan Provincial People’s Hospital. We included and excluded patients according to the following criteria. Inclusion criteria were patients with basic disease causing heart failure including hypertension, coronary heart disease, diabetes, chronic obstructive pulmonary disease, various primary cardiomyopathies, and various secondary cardiomyopathies. Exclusion criteria were acute myocardial infarction, acute renal tubular nephritis and other acute kidney disease, severe acute infection, severe acute organ failure, malignant tumour, and failure to complete data collection. Informed written consent was provided by all study participants. All patients who gave their consent to participate were included in this study, following the principles of the Declaration of Helsinki and local regulations for the protection of medical data. The study protocol was approved by the Ethics Committee of Sichuan Provincial People’s Hospital (ethical approval document No. 382, 2020).

### Data Collection

The basic information of the population included age; sex; basic disease data including medical history and frailty score at admission; blood test data including serum creatinine, estimated glomerular filtration rate, and B-type brain natriuretic peptide levels; and echocardiography data including left atrial diameter, left ventricular end diastolic diameter, left ventricular posterior wall thickness, interventricular septal thickness, right ventricular, right atrial diameter, pulmonary artery velocity, pulmonary valve pressure gradient, tricuspid regurgitation velocity, tricuspid regurgitation pressure gradient, aortic valve velocity, aortic valve pressure gradient, early diastolic mitral inflow peak velocity (E peak), late diastolic mitral inflow peak velocity (A peak), early diastolic Doppler spectrum of mitral valve (e peak), late diastolic Doppler spectrum of mitral valve (a peak), left ventricular ejection fraction, left ventricular fractional shortening, and tricuspid annular plane systolic excursion. Follow-up data included whether patients died within 1 year of discharge, whether and how often they were readmitted to hospital, and whether they were readmitted to hospital because of heart failure.

Basic data and disease data were based on admission records, and diagnosis of disease was based on the guideline diagnostic criteria of various disciplines. According to clinical work guidelines developed by the International Conference of Frailty and Sarcopenia Research, we used the FRAIL scale to assess frailty.[9] The blood test data were provided by the Clinical Laboratory of Sichuan Provincial People’s Hospital. Echocardiography data were measured and recorded by professional echocardiographic personnel in the Ultrasonography Department of Sichuan Provincial People’s Hospital, who were trained according to the American Society of Echocardiography guidelines.[10] Follow-up data were obtained in monthly outpatient visits and telephone interviews performed by trained staff.

### Statistical Analysis

IBM SPSS 25.0 was used for statistical analysis of the data. Data are expressed as the mean with standard deviation (mean±SD) for normally distributed variables, and as the median with interquartile range (median [Q25, Q75]) for non-normally distributed data. Categorical data are expressed as numbers and percentages. Group differences were evaluated using Student’s t-test or Kruskal–Wallis test for continuous variables and the chi-squared test or Fisher’s exact test for categorical variables. Event-free survival curves were constructed using the Kaplan–Meier survival method and were compared using log-rank statistics. The association of baseline frailty and other variables with the time to all-cause mortality, all-cause admission, and heart failure-related readmission are summarized with hazard ratios and their 95% confidence intervals obtained from Cox regression. In all statistical analyses, statistical tests were two-sided, and P<0.05 was considered significant.

## Results

A total of 226 people aged ≥80 years were enrolled. Because seven patients were lost to follow-up, 219 people were included in the final analysis. Using the FRAIL scale score, the population was divided into three groups; those with a score of 0 were classified in the non-frailty group, those with a score of 1–2 in the pre-frailty group, and those with a score ≥3 in the frailty group. The baseline characteristics are shown in Table 1.

**Table 1.**
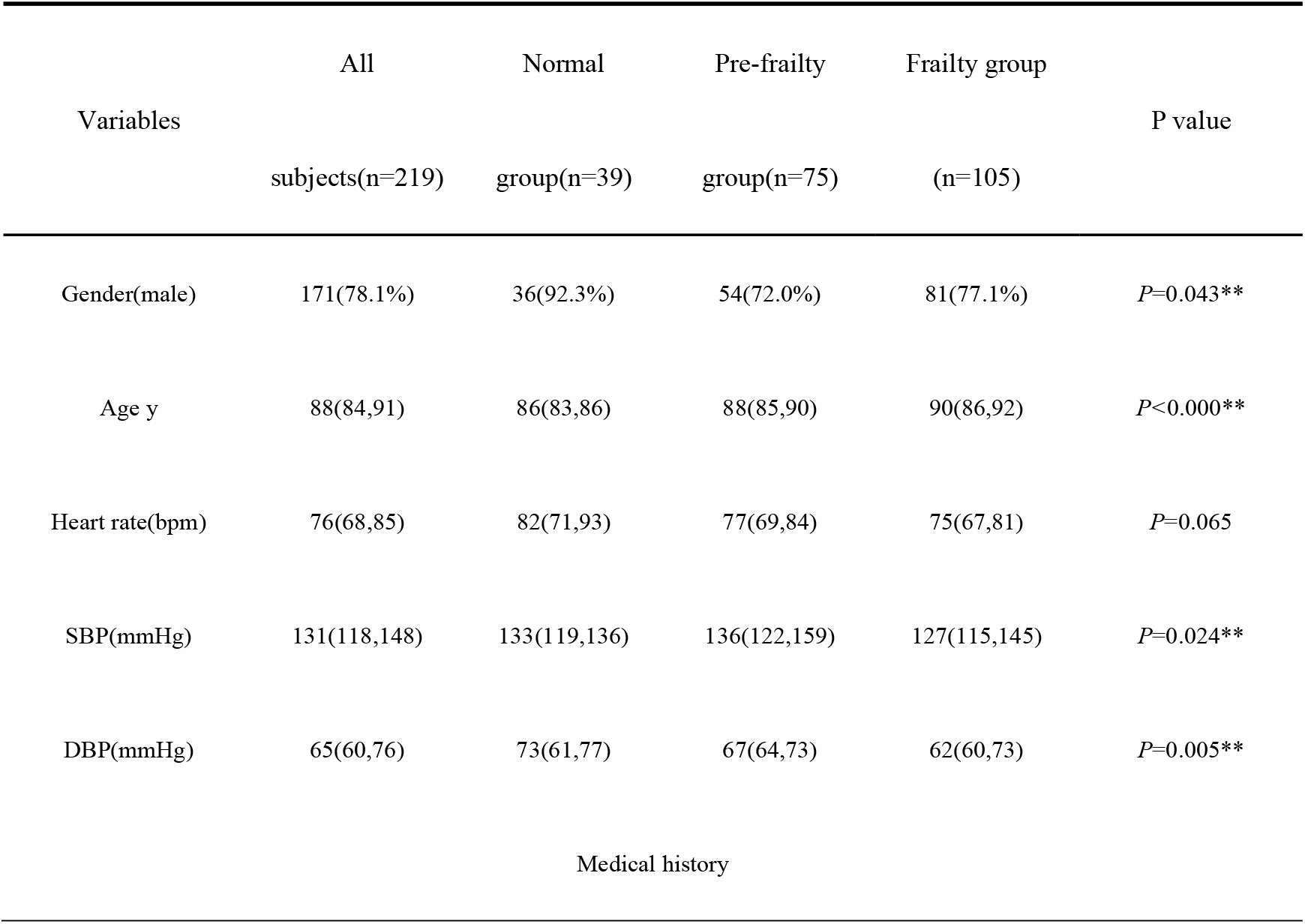
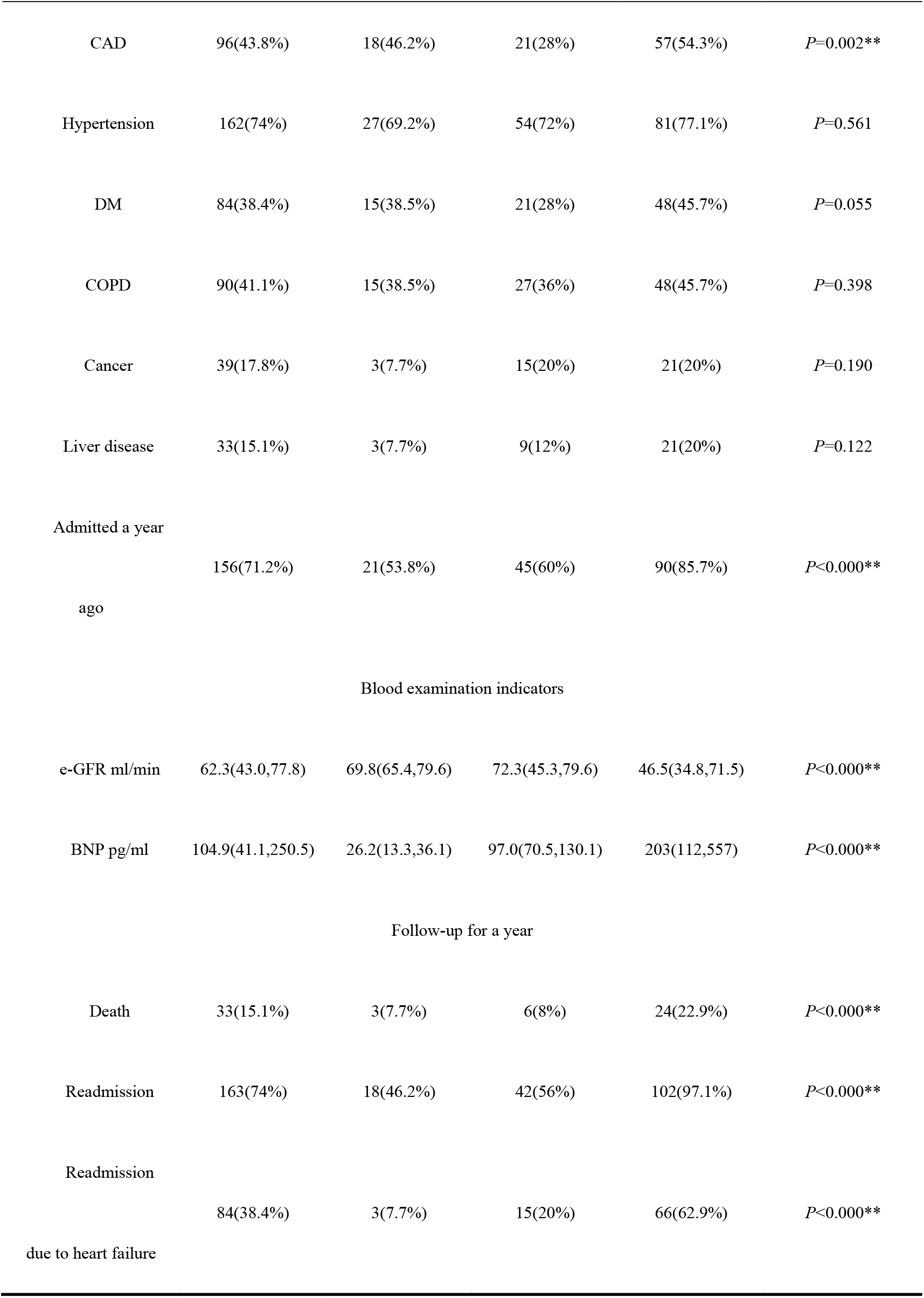
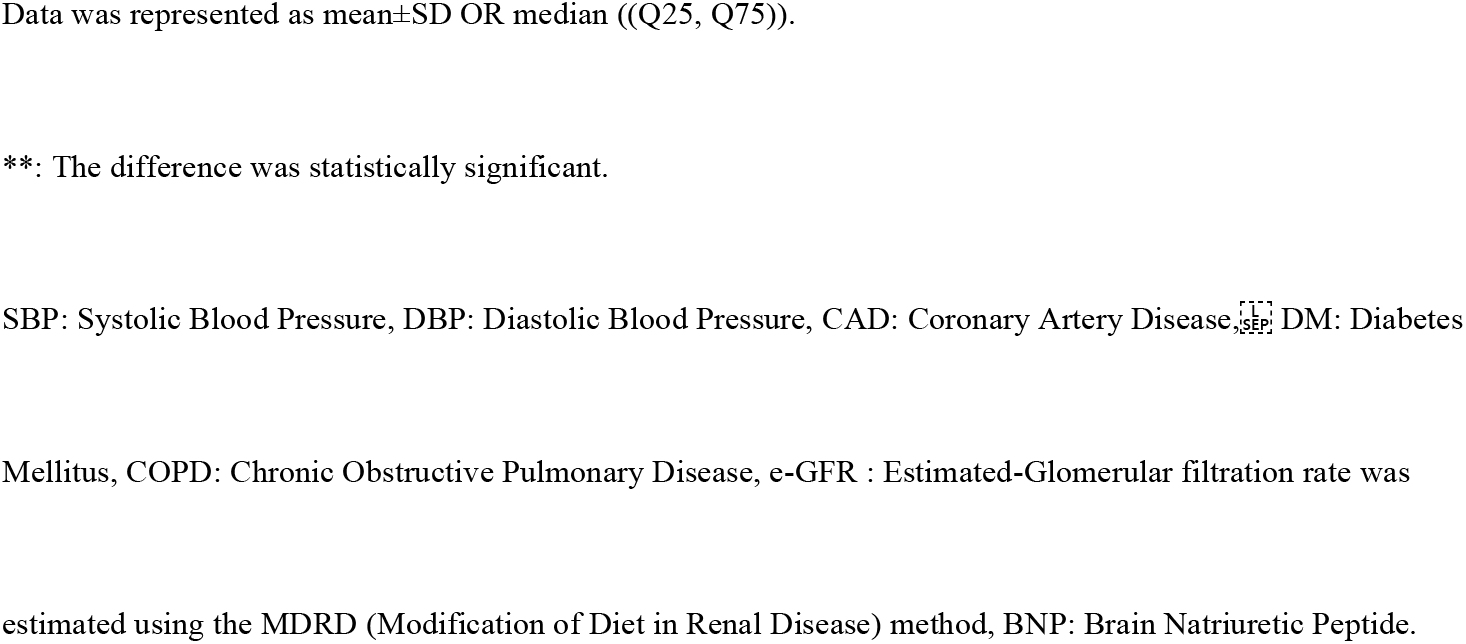
Baseline demographic and clinical characteristics

### Echocardiographic Data Analysis

There were significant differences in cardiac structure between all groups. Diameters in the frailty group were significantly larger than those in the other groups (left atrial diameter: 40 [39, 45], P<0.000; left ventricular end diastolic diameter: 46 [43, 50], P=0.001; right atrial diameter: 51.0 (45, 60), P=0.000; right ventricular diameter: 21 [20, 23], P=0.000). The left ventricular posterior wall and interventricular septum were also thicker in the frailty group (intraventricular septal thickness: 10 [10, 11], P<0.000; left ventricular posterior wall thickness: 10 [9, 11], P=0.002). In terms of valve lesions, the frailty group had more valve damage than the normal group (3 [2, 3], P<0.000); The degree of valve damage in each group gradually increased with increasing FRAIL score (P<0.000). In terms of heart function, left ventricular ejection fraction in the frailty group was lower than that in the other groups (0.65 [0.62, 0.67], P=0.01). Tricuspid annular plane systolic excursion decreased gradually from non-frailty to frailty group (P<0.000; Table 2). The proportion of patients with right heart failure in the frailty group was significantly higher (16.3% vs. left heart failure 7%, P=0.031) (See the attached Fig. A).

**Table 2.**
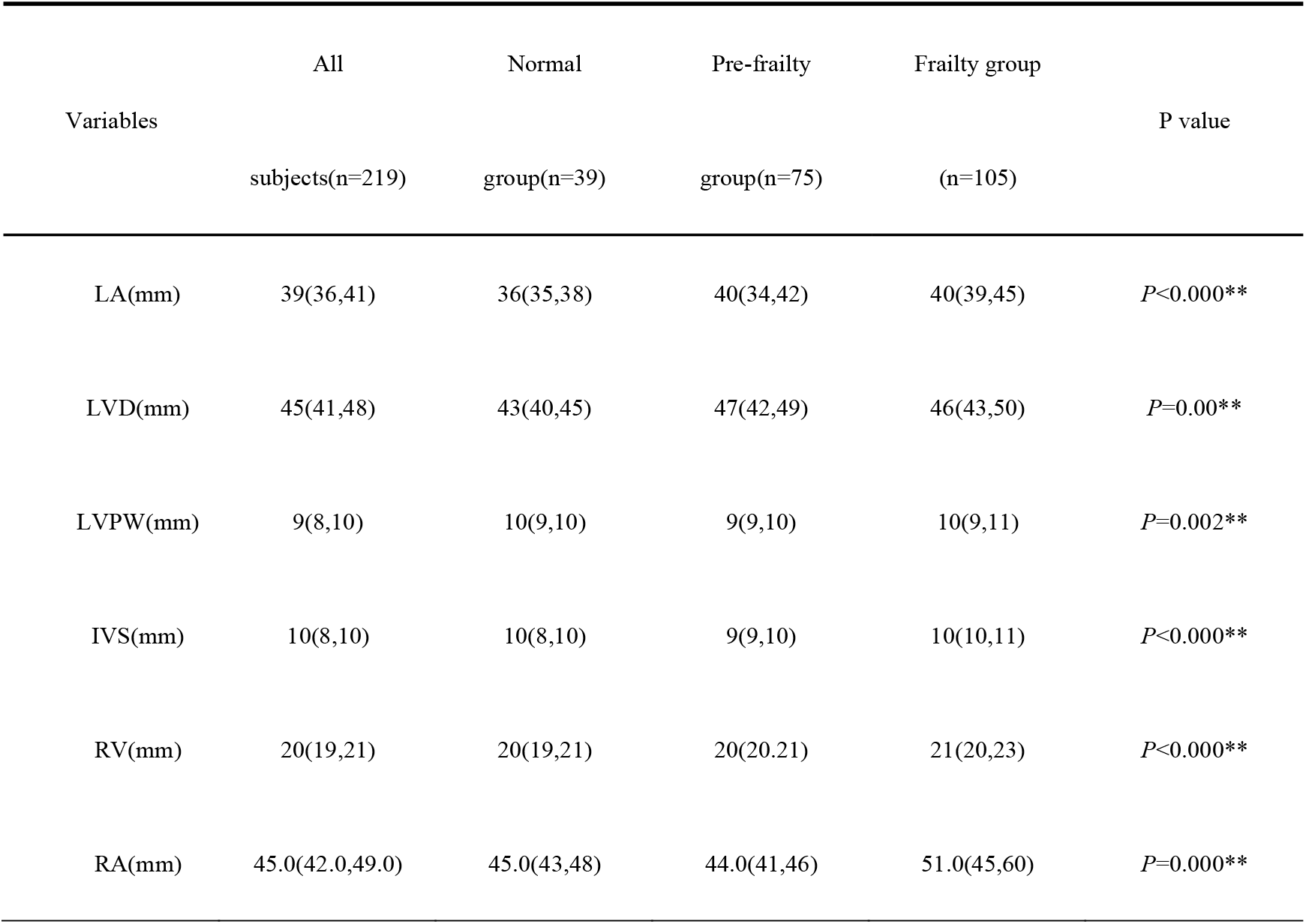
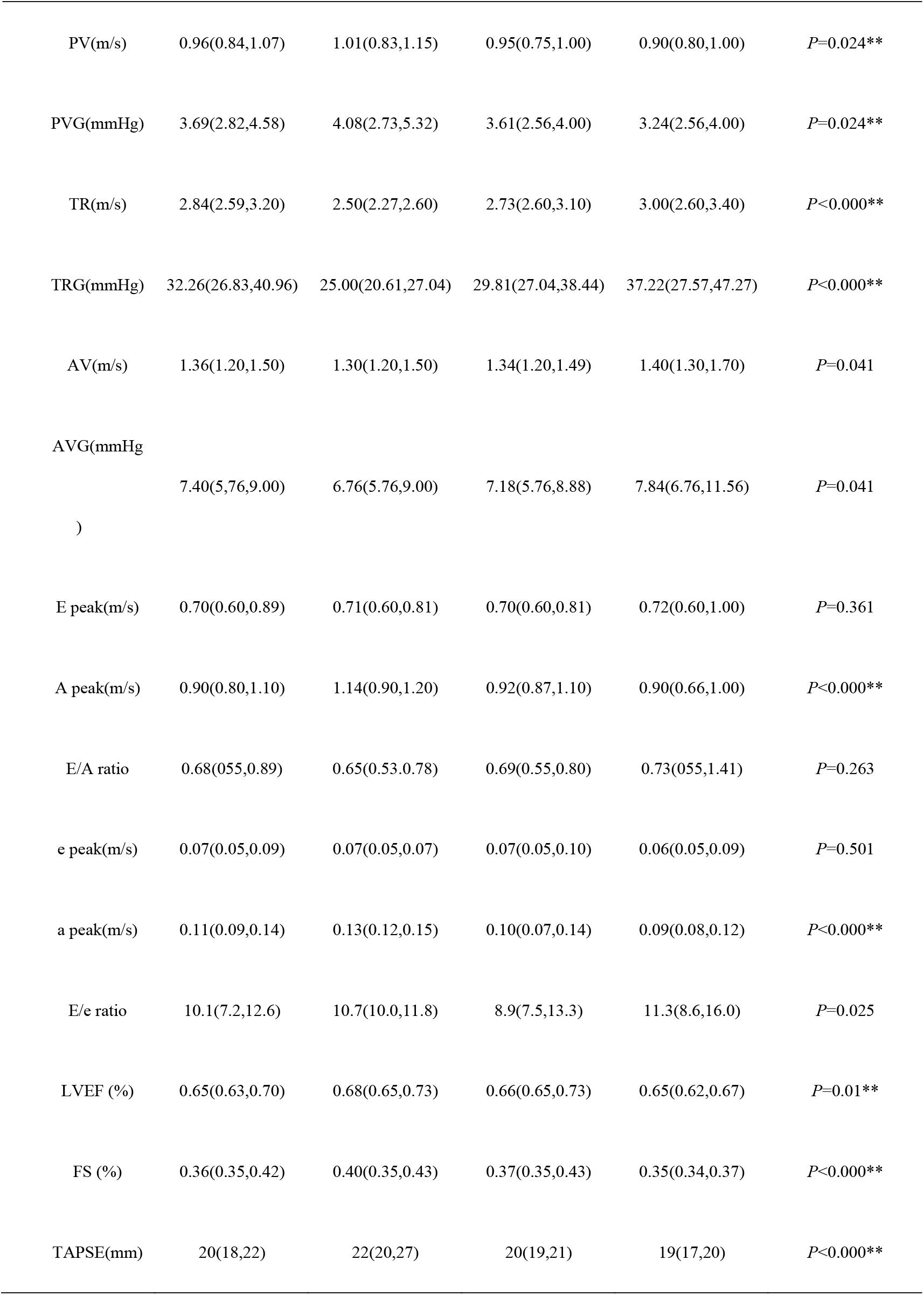
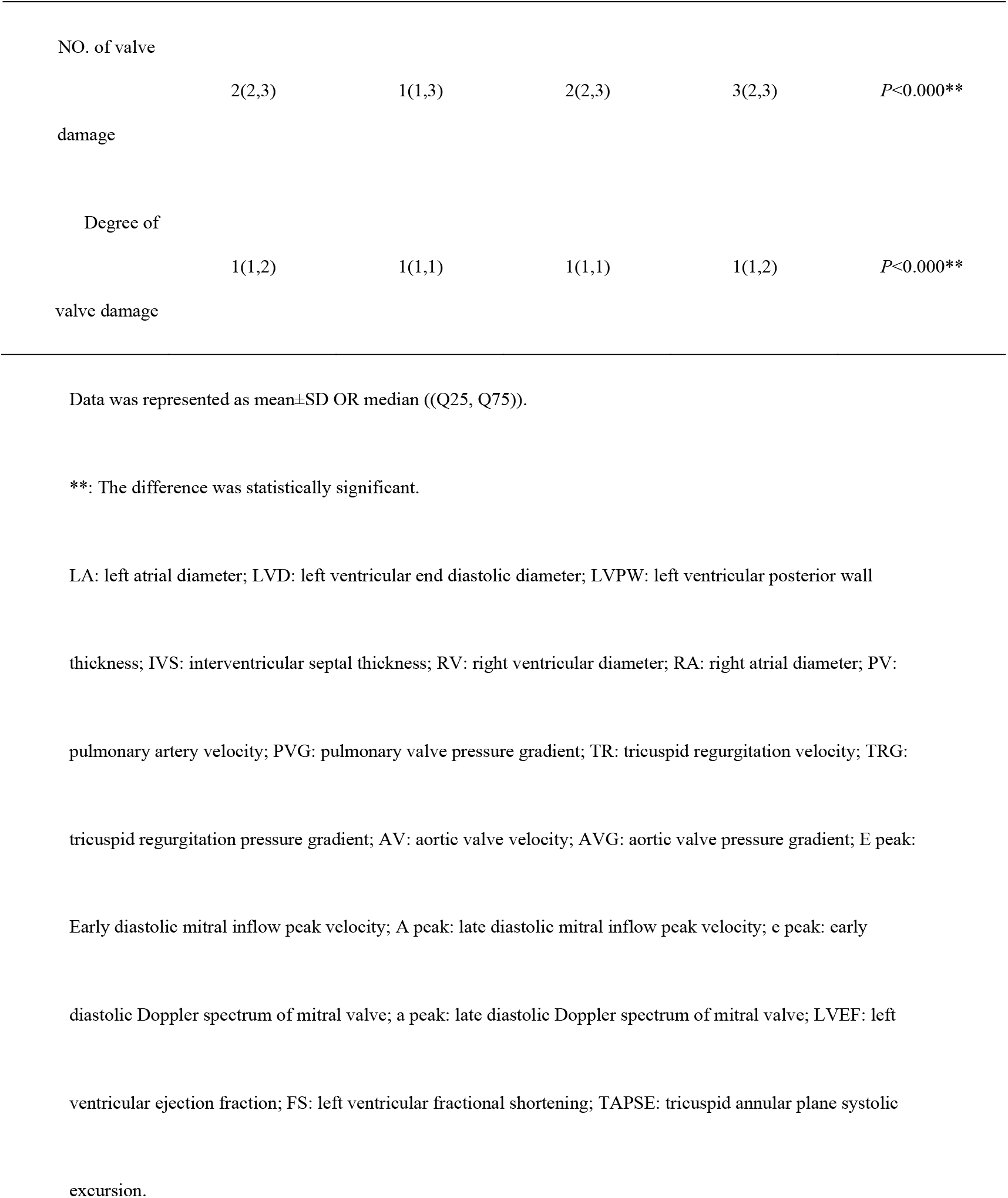
Characteristics of echocardiography data

### Kaplan–Meier Analysis

Among the 219 senile patients, 33 (15.1%) died, 162 (74%) were readmitted, and 84 (38.4%) were admitted for heart failure within 1 year. To explore the differences in one-year mortality, all-cause readmission rate, and heart failure-related readmission rate under different clinical statuses, we divided the population into five groups for survival analysis according to whether there was frailty or heart failure: frailty with left heart failure, frailty with right heart failure, frailty without heart failure, heart failure without frailty, and other (See the attached Fig. B). Kaplan–Meier analysis was performed based on the above grouping, and the results are shown in Figure 1.

Significant differences were observed in the influence of each group on three events. Frailty with right heart failure had a significantly greater effect on all three events than frailty with left heart failure. We also analysed the influence of frailty, left heart failure, and right heart failure on the three follow-up target events, and the results are shown in Figure 2. We found that frailty had significant effects on 1-year all-cause mortality, all-cause readmission rate, and heart failure-related readmission rate; the risk of each event was significantly higher in the frailty group than in the other groups. The rate of the three events in patients with left heart failure showed a nonsignificant increase. Right heart failure was related to significantly increased 1-year all-cause admissions and heart failure-related admissions, but not 1-year all-cause mortality.

**Figure 2:**
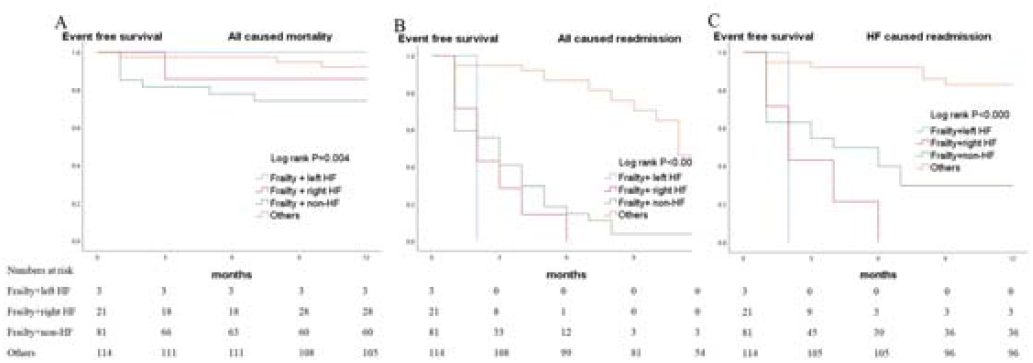
Kaplan-Meier analysis based by frailty and heart failure combination 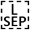 A: All caused mortality;B: All caused readmission; C: HF caused readmission 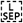: HF: heart failure

**Figure 3:**
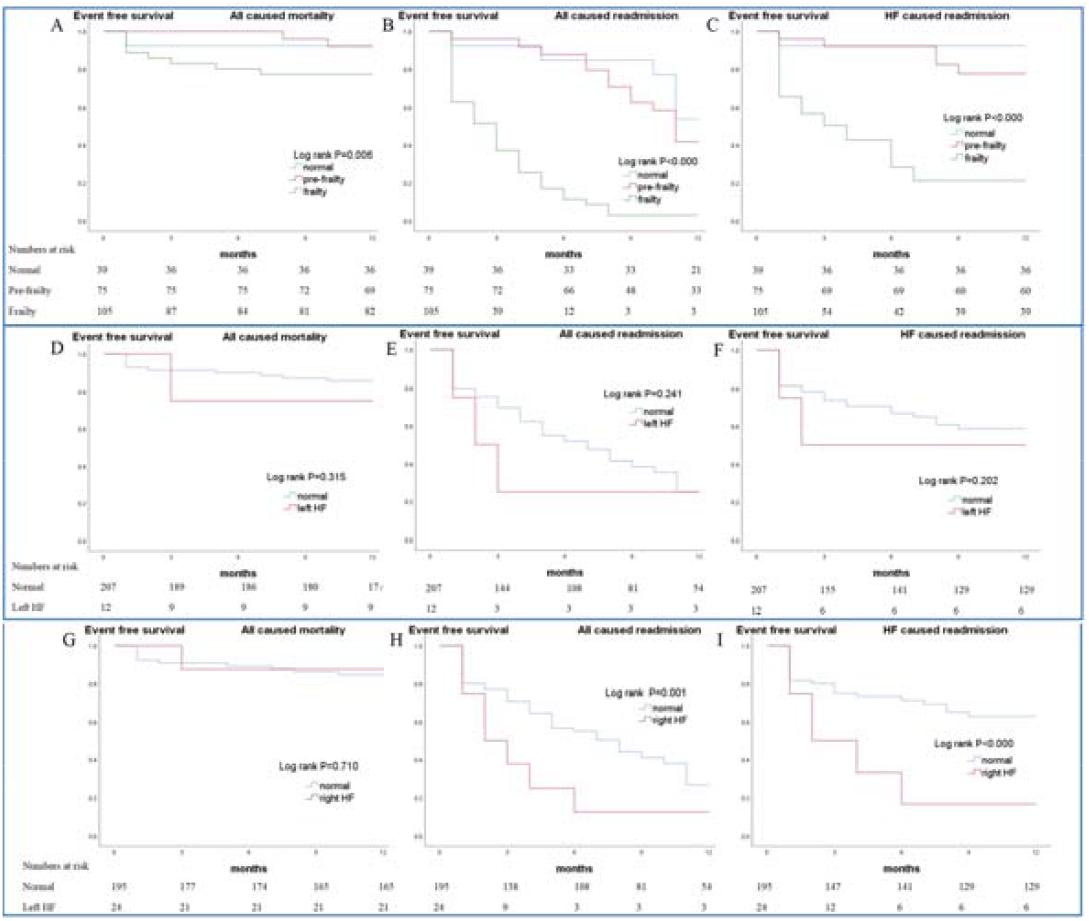
Kaplan-Meier analysis based by frailty, left heart failure and right heart failure. A: All caused mortality by frailty group. 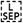B: All caused readmission by frailty group. C: HF caused readmission by frailty group 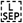D: All caused mortality by left heart failure group. 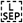 E: All caused readmission by left heart failure group. 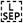 F: HF caused readmission by left heart failure group. G: All caused mortality by right heart failure group. 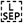 H: All caused readmission by right heart failure group. 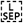 I: HF caused readmission by right heart failure group. HF: heart failure

### Cox Regression Analysis

We performed a Cox equal-ratio risk model analysis for frailty, left heart failure, and right heart failure and calculated hazard ratio values after adjusting for the three factors. Three adjusted models were established, which were as follows: Model 1, adjusted for sex, age, systolic blood pressure, diastolic blood pressure, coronary heart disease, diabetes mellitus, hospital admittance in the previous year, and estimated glomerular filtration rate; Model 2, Model 1 + left atrial diameter, left ventricular end diastolic diameter, left ventricular posterior wall thickness, intraventricular septal thickness, right ventricular diameter, right atrial diameter, number of damaged valves, and degree of valve damage; and Model 3, Model 2 + left ventricular ejection fraction, tricuspid annular plane systolic excursion, and brain natriuretic peptide level. The analysis results are listed in Table 3. The effects of frailty on 1-year mortality, readmission, and heart failure-related readmission were statistically significant in almost all adjusted models. This effect became more significant with an increase in adjusting variables. Left heart failure had an overall effect of increasing the risk of the three events; however, the results were inconsistent across the different adjusted models and not significant without adjustment or after final adjustment (Model 3). There was no significant effect of right heart failure on mortality regardless of adjustment status. However, in the unadjusted analysis, right heart failure was related to increased risk of 1-year readmission and heart failure-related readmission (2.05 [1.29–3.27], P=0.002 and 2.94 [1.73–4.99], P<0.000, respectively); this significance disappeared after adjustment.

**Table 3.**
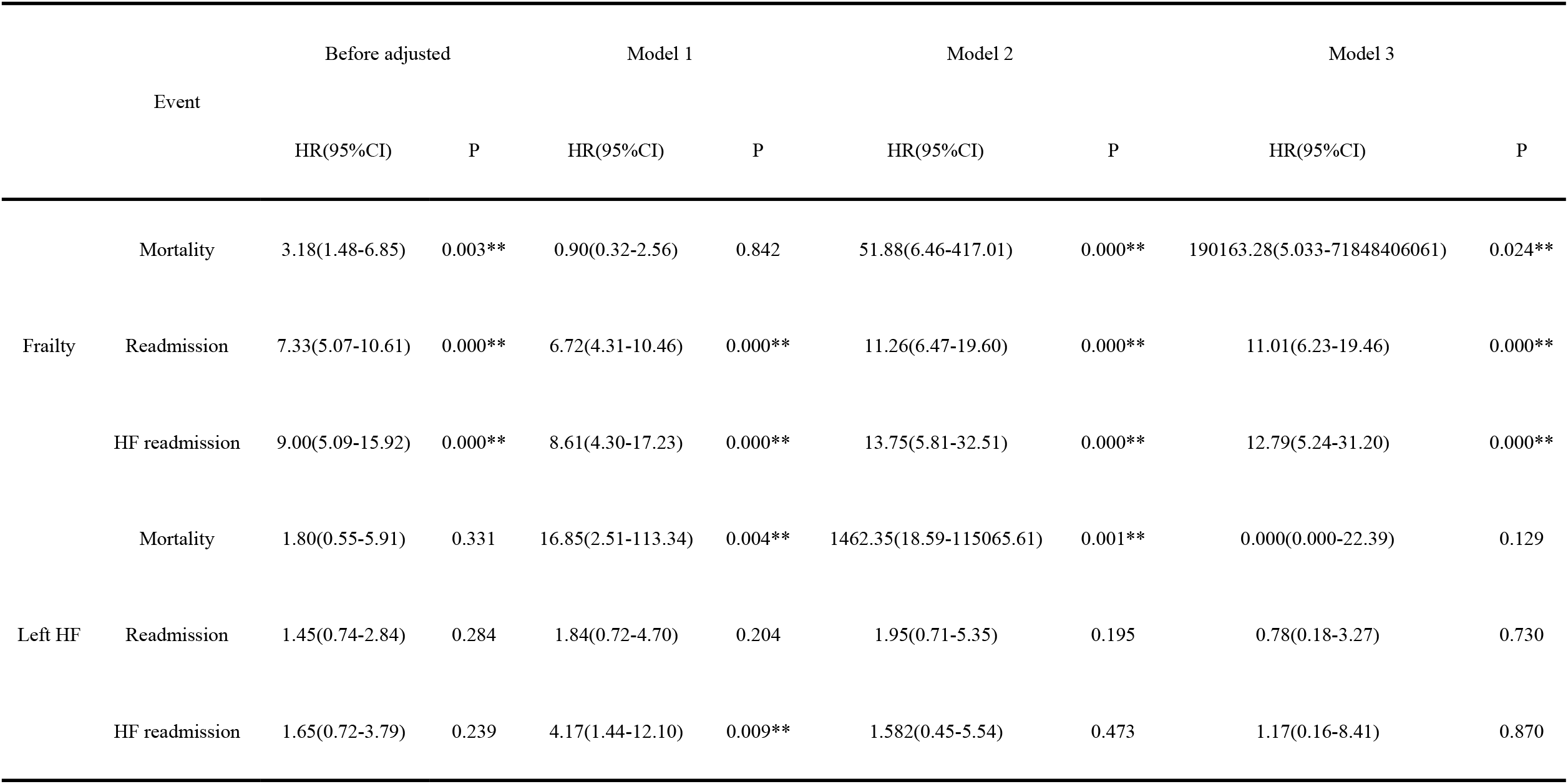
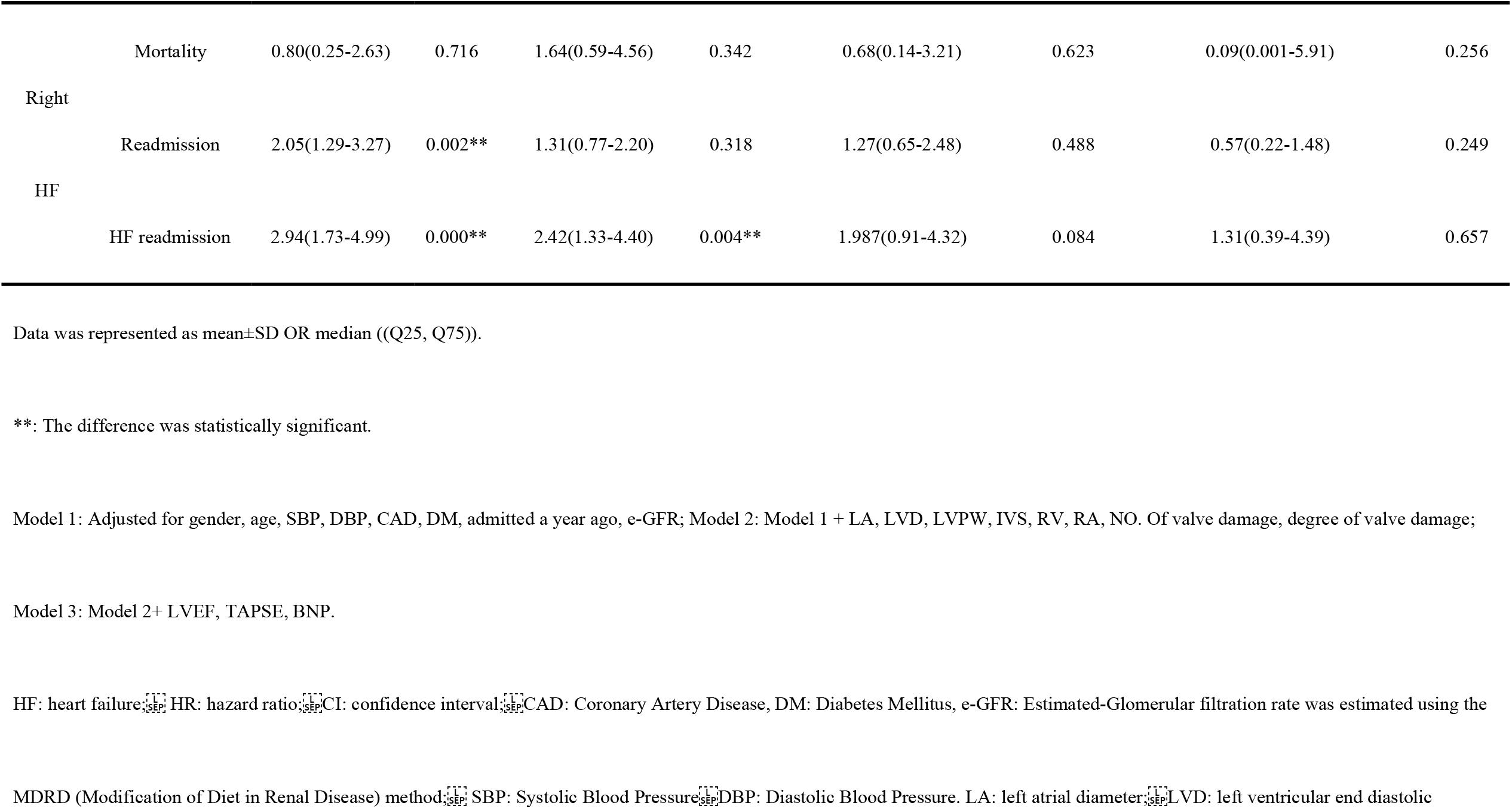
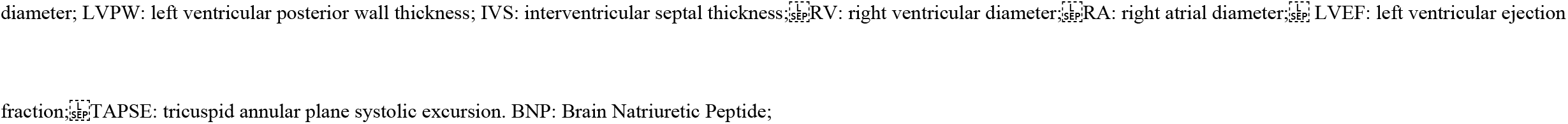
Cox-regression analysis

### Discussions

This study was performed in elderly patients to examine the influence of frailty on biventricular function and explore the difference in prognosis of patients with different clinical statuses. Our main findings were as follows: frailty was more likely to be associated with right heart failure than left heart failure; the 1-year all-cause mortality, all-cause readmission, and heart failure-related readmission rates of frailty patients with right heart failure were significantly increased; and frailty was an important predictor of all-cause death, all-cause readmission, and heart failure-related readmission.

## Discussion of Basic Data

### Frailty and cardiac structure disorders

The effect of frailty on the structure of the heart has been relatively well studied both at the level of the heart and the myocardia. heart failure is accelerated with increasing frailty. A multi-omics study demonstrated that changes in atrial structure and function significantly correlated with frailty index, and the more obvious the frailty was, the greater the changes in atrial structure and function. This change conforms to the hypothesis that the extracellular matrix is regulated by matrix metalloproteinases and tissue inhibitors of metalloproteinases, which determine atrial fibrosis.[24] A Japanese study of elderly people reported that the left atrial diameter in patients with frailty increased significantly,[23] which was consistent with this mechanism from a macro perspective. An animal study found a positive correlation between heart weight and frailty after adjusting for age; at the same time, the researchers also found a positive correlation between ventricular myocyte width and frailty, an association that did not exist when analysed with respect to age.[25] Our study confirmed that the frailty group had larger heart chambers and thicker ventricular walls, consistent with previous studies.

### Frailty is more likely to be associated with right heart failure

Current research data on left and right ventricles suggested that left and right ventricular failure often occurred together;[11] however, even without considering frailty, left and right ventricular failure was relatively independent. Valentova et al. reported that cardiac cachexia was associated with right heart failure and elevated central venous pressure, but not with left ventricular structure or function.[12] In our study, frailty with right heart failure was more than twice as common as frailty with left heart failure, which is consistent with previous studies.[13] The reason for this phenomenon may be that the left and right ventricles originate from different progenitor cells.[14] Compared with the left ventricle, the right ventricle has a thinner wall and poorer resistance to deformation; therefore, it is more prone to deformation and systolic or diastolic dysfunction when subjected to volume overload.[15] Of course, further research is needed to explore deeper causes and make accurate comparisons.

### Discussion of Follow-up Data

The all-cause mortality rate in our population after 1 year of follow-up was 15.1%. In a study of cardiac resynchronization therapy patients with an average age of 79 years, the all-cause mortality rate of 151 patients was 10% after 9 months of follow-up;[26] thus, the observed all-cause mortality rate was close to expected considering the high average age of our study population (88 years). During the 1-year follow-up of our study, 74% (162) of patients were readmitted and 38.4% (84) were admitted for heart failure. A report published in 2020 based on United States Medicare Big Data noted an increase in 30-day and 90-day all-cause readmission rates for patients with heart failure from 2010 to 2017, with a 90-day all-cause readmission rate of 34.6% in 2017. The mean age of patients in that study was 71.5 years.[16] In the present study, the population of the Elderly Cadre Department has the characteristics of adequate economic capacity, sufficient family care, high average age, high incidence of clinical complications, and high follow-up compliance, which can explain the higher readmission rate and heart failure-related readmission rate in our population. Frailty affects mortality, readmission, and heart failure-related readmission rates In the cardiovascular field, frailty increases the risk of death and readmission because of coronary heart disease,[17] cardiac interventional surgery,[18] and other diseases. A 2021 study confirmed frailty as an independent predictor of mortality and readmission in patients with heart failure with preserved ejection fraction.[30] In our study, frailty had a significant effect on 1-year mortality, readmission rates, and increased rates of readmission because of to heart failure in senile hospitalized patients, consistent with previous findings, regardless of adjusting for the effect of heart failure.

### Right heart failure affects mortality, readmission, and heart failure-related readmission rates

The influence of right heart failure on clinical prognosis has been studied. In 2007, tricuspid annular plane systolic excursion was reported to be associated with increased mortality in patients with heart failure.[19] In a study of decompensated heart failure, it was found that low tricuspid annular plane systolic excursion was associated with more mortality risk factors, which may have weakened the effect of tricuspid annular plane systolic excursion on mortality after weighted analysis.[20] As our study targeted elderly patients with heart failure with a large number of comorbidities, no significant effect was observed when exploring the influence of right heart failure on mortality, which may be due to the high incidence of frailty in our population. After the effects of frailty were combined, the impact of right heart failure on mortality was weakened. Frailty is an inevitable problem in the elderly, especially in the senile. Therefore, this result may somehow reflect the real-world situation; however, it still needs confirmation in a large sample study. Our data suggest that right heart failure increases the risk of readmission and heart failure-related readmission. It has been reported that non-cardiovascular factors account for a higher hospitalization rate in patients with heart failure with preserved ejection fraction.[21] In addition, the symptoms and signs of patients with right heart failure, such as lower limb oedema, gastrointestinal congestion, and loss of appetite, are obvious and are easy for patients to notice, which is also an important reason for patients to be hospitalized.

### Left heart failure affects mortality, readmission, and heart failure-related readmission rates

Left heart failure’s impact on mortality, readmission, and heart failure-related readmission rates is significant.[25] In 2017, Kubala et al. found that severe left ventricular systolic dysfunction increased the risk of death patients with heart failure.[26] In our data, there was no significant effect of left heart failure on mortality, readmission, or heart failure-related readmission rates. The reasons were hypothesized to be as follows: 1) our sample was small, and the sample of patients with left heart failure was even smaller; and 2) left heart failure was defined based on reduced left ventricular ejection fraction in our study. The reported incidence of this type of heart failure in the elderly is not as high as that of heart failure with preserved ejection fraction, which may be due to the shorter overall survival of patients with left heart failure.[27]

Combination analysis: frailty + left heart failure, frailty + right heart failure

We found that the ability of frailty to influence all three prognostic events was preserved when combined with right heart failure, while this effect was obscured when frailty was combined with left heart failure. At present, there are few studies that compare left and right ventricular failure, and this comparison is especially rare in a frail population. We defined left heart failure as an ejection fraction below 50%, i.e., heart failure with reduced ejection fraction. Right heart failure, which is often closely related to reduced left ventricular diastolic function, has been shown to be an important component of heart failure with preserved ejection fraction.[28] It has been reported that the incidence of heart failure with preserved ejection fraction, especially in the elderly, is higher than that of other types of heart failure.[29] The average age of our study population was 88 years old, which partly explains why we had fewer cases of frailty with left heart failure. Therefore, we conclude that frailty with right heart failure was a strong predictor of mortality, readmission, and heart failure-related readmission rates, but comparisons with frailty combined with left heart failure should be cautious.

### Limitations

The present study had several limitations. First, the study population was small and from a single centre, which may affect the strength of our conclusions. Second, in our study population, there were only a few subjects in the frailty group with left heart failure, although this was somehow related to the characteristics of people with a high incidence of left heart failure. We need a larger sample size to expand the scope of the study in the future to reinforce our conclusions. In addition, we used only one frailty assessment protocol to determine frailty, which may result in a decrease in the comparability of our results with previously published literature using other or multiple scales.

## Data Availability

All relevant data are within the paper.

## Conclusions

We found that frailty was more likely to be associated with right heart failure than left heart failure; 1-year all-cause mortality, all-cause readmission, and heart failure-related readmission rates were significantly higher in patients with frailty and right heart failure; and frailty was a significant predictor of all-cause death, all-cause readmission, and heart failure-related readmission. These findings highlight the effect of frailty on right heart failure, the importance of frailty with right heart failure in clinical practice, and provide new perspectives for further optimization of clinical management of senile patients in the future.

## References

[1] Matsue Y, Kamiya K, Saito H, Saito K, Ogasahara Y, Maekawa E, et al. Prevalence and prognostic impact of the coexistence of multiple frailty domains in elderly patients with heart failure: the FRAGILE-HF cohort study. Eur J Heart Fail. 2020;22(11):2112–9.

[2] Uchmanowicz I, Lee CS, Vitale C, Manulik S, Denfeld QE, Uchmanowicz B, et al. Frailty and the risk of all-cause mortality and hospitalization in chronic heart failure: a meta-analysis. ESC Heart Fail. 2020 Sep 21;7(6):3427–37.

[3] McDonagh J, Ferguson C, Newton PJ. Frailty Assessment in Heart Failure: an Overview of the Multi-domain Approach. Curr Heart Fail Rep. 2018 Feb;15(1):17–23.

[4] Farmakis D, Thodi M, Elpidoforou M, Filippatos G. Assessing frailty in heart failure. Eur J Heart Fail. 2020 Nov;22(11):2134–7.

[5] Ponikowski P, Voors AA, Anker SD, Bueno H, Cleland JG, Coats AJ, et al. Authors/Task Force Members; Document Reviewers. 2016 ESC Guidelines for the diagnosis and treatment of acute and chronic heart failure: The Task Force for the diagnosis and treatment of acute and chronic heart failure of the European Society of Cardiology (ESC). Developed with the special contribution of the Heart Failure Association (HFA) of the ESC. Eur J Heart Fail. 2016 Aug;18(8):891–975.

[6] Sze S, Pellicori P, Zhang J, Weston J, Clark AL. Identification of Frailty in Chronic Heart Failure. JACC Heart Fail. 2019 Apr;7(4):291–302.

[7] Valdiviesso R, Azevedo LF, Moreira E, Ataíde R, Martins S, Fernandes L, et al. Frailty phenotype and associated nutritional factors in a sample of Portuguese outpatients with heart failure. Nutr Metab Cardiovasc Dis. 2021 Jul 22;31(8):2391–7.

[8] Ferrari MW, Schulze PC, Kretzschmar D. Acute right heart failure: future perspective with the PERKAT RV pulsatile right ventricular support device. Ther Adv Cardiovasc Dis. 2020 Jan-Dec;14:1753944719895902.

[9] Dent E, Morley JE, Cruz-Jentoft AJ, Woodhouse L, Rodríguez-Mañas L, Fried LP, et al. Physical Frailty: ICFSR International Clinical Practice Guidelines for Identification and Management. J Nutr Health Aging. 2019;23(9):771–87.

[10] Lang RM, Badano LP, Mor-Avi V, Afilalo J, Armstrong A, Ernande L, et al. Recommendations for cardiac chamber quantification by echocardiography in adults: an update from the American Society of Echocardiography and the European Association of Cardiovascular Imaging. J Am Soc Echocardiogr. 2015 Jan;28(1):1-39.e14.

[11] Konstam MA, Kiernan MS, Bernstein D, Bozkurt B, Jacob M, Kapur NK, et al. American Heart Association Council on Clinical Cardiology; Council on Cardiovascular Disease in the Young; and Council on Cardiovascular Surgery and Anesthesia. Evaluation and Management of Right-Sided Heart Failure: A Scientific Statement From the American Heart Association. Circulation. 2018 May 15;137(20):e578–e622.

[12] Valentova M, von Haehling S, Bauditz J, Doehner W, Ebner N, Bekfani T, et al. Intestinal congestion and right ventricular dysfunction: a link with appetite loss, inflammation, and cachexia in chronic heart failure. Eur Heart J. 2016 Jun 1;37(21):1684–91.

[13] Pandey A, Gilbert O, Kitzman DW. Physical frailty in older patients with acute heart failure: From risk marker to modifiable treatment target. J Am Geriatr Soc. 2021 Sep;69(9):2451–4.

[14] Shults NV, Melnyk O, Suzuki DI, Suzuki YJ. Redox Biology of Right-Sided Heart Failure. Antioxidants (Basel). 2018 Aug 8;7(8):106.

[15] Thandavarayan RA, Chitturi KR, Guha A. Pathophysiology of Acute and Chronic Right Heart Failure. Cardiol Clin. 2020 May;38(2):149–60.

[16] Khan MS, Sreenivasan J, Lateef N, Abougergi MS, Greene SJ, Ahmad T, et al. Trends in 30- and 90-Day Readmission Rates for Heart Failure. Circ Heart Fail. 2021 Apr;14(4):e008335.

[17] Zhang S, Meng H, Chen Q, Wang X, Zou J, Hao Q, Yang M, Wu J. Is frailty a prognostic factor for adverse outcomes in older patients with acute coronary syndrome? Aging Clin Exp Res. 2020 Aug;32(8):1435–42.

[18] Anand A, Harley C, Visvanathan A, Shah ASV, Cowell J, MacLullich A, et al. The relationship between preoperative frailty and outcomes following transcatheter aortic valve implantation: a systematic review and meta-analysis. Eur Heart J Qual Care Clin Outcomes. 2017 Apr 1;3(2):123–32.

[19] Kjaergaard J, Akkan D, Iversen KK, Køber L, Torp-Pedersen C, Hassager C. Right ventricular dysfunction as an independent predictor of short- and long-term mortality in patients with heart failure. Eur J Heart Fail. 2007 Jun-Jul;9(6-7):610–6.

[20] Scrutinio D, Catanzaro R, Santoro D, Ammirati E, Passantino A, Oliva F, et al. Tricuspid Annular Plane Systolic Excursion in Acute Decompensated Heart Failure: Relevance for Risk Stratification. Can J Cardiol. 2016 Aug;32(8):963–9.

[21] Gerber Y, Weston SA, Redfield MM, Chamberlain AM, Manemann SM, Jiang R, et al. A contemporary appraisal of the heart failure epidemic in Olmsted County, Minnesota, 2000 to 2010. JAMA Intern Med. 2015 Jun;175(6):996–1004.

[22] Kusunose K, Okushi Y, Yamada H, Nishio S, Torii Y, Hirata Y, et al. Prognostic Value of Frailty and Diastolic Dysfunction in Elderly Patients. Circ J. 2018 Jul 25;82(8):2103–10.

[23] Jansen HJ, Moghtadaei M, Mackasey M, Rafferty SA, Bogachev O, Sapp JL, et al. Atrial structure, function and arrhythmogenesis in aged and frail mice. Sci Rep. 2017 Mar 14;7:44336.

[24] Feridooni HA, Kane AE, Ayaz O, Boroumandi A, Polidovitch N, Tsushima RG, Rose RA, Howlett SE. The impact of age and frailty on ventricular structure and function in C57BL/6J mice. J Physiol. 2017 Jun 15;595(12):3721–42.

[25] Pocock SJ, Ariti CA, McMurray JJ, Maggioni A, Køber L, Squire IB, et al. Meta-Analysis Global Group in Chronic Heart Failure. Predicting survival in heart failure: a risk score based on 39 372 patients from 30 studies. Eur Heart J. 2013 May;34(19):1404–13.

[26] Kubala M, Guédon-Moreau L, Anselme F, Klug D, Bertaina G, Traullé S, et al. Utility of Frailty Assessment for Elderly Patients Undergoing Cardiac Resynchronization Therapy. JACC Clin Electrophysiol. 2017 Dec 26;3(13):1523–33.

[27] Alter DA, Ko DT, Tu JV, Stukel TA, Lee DS, Laupacis A, et al. The average lifespan of patients discharged from hospital with heart failure. J Gen Intern Med. 2012 Sep;27(9):1171–9.

[28] Kanagala P, Arnold JR, Singh A, Khan JN, Gulsin GS, Gupta P, et al. Prevalence of right ventricular dysfunction and prognostic significance in heart failure with preserved ejection fraction. Int J Cardiovasc Imaging. 2021 Jan;37(1):255–66.

[29] Cho DH, Yoo BS. Current Prevalence, Incidence, and Outcomes of Heart Failure with Preserved Ejection Fraction. Heart Fail Clin. 2021 Jul;17(3):315–26.

[30] Goyal P, Yum B, Navid P, Chen L, Kim DH, Roh J, Jaeger BC, Levitan EB. Frailty and Post-hospitalization Outcomes in Patients With Heart Failure With Preserved Ejection Fraction. Am J Cardiol. 2021 Jun 1;148:84–93.

